# Seroresponse to Inactivated and Recombinant Influenza Vaccines among Maintenance Hemodialysis Patients

**DOI:** 10.1101/2021.12.17.21267999

**Authors:** Harold J. Manley, Eduardo K. Lacson, Gideon Aweh, Daniel E. Weiner, Dana C. Miskulin, Caroline M. Hsu, Toros Kapoian, Mary S. Hayney, Klemens B. Meyer, Doug S. Johnson

## Abstract

**Rationale & Objectives:** High dose influenza vaccine provides better protection against influenza infection in older adults than standard dose vaccine. We compared vaccine seroresponse among hemodialysis patients over 4 months after high dose inactivated (HD-IIV3), standard dose inactivated (SD-IIV4) or recombinant (RIV4) influenza vaccine.

**Study Design:** Prospective observational study.

**Setting & Participants:** Patients at four hemodialysis clinics who received influenza vaccination. Hemagglutination inhibition titers were measured at baseline and at 1, 2, 3 and 4 months following vaccination with HD-IIV3, SD-IIV4 or RIV4 influenza vaccine.

**Outcome:** The primary outcome was seroprotection rates at ≥1:40 and ≥1:160 (which correspond to antibody levels providing protection from infection in about 50% and 95% immunocompetent individuals, respectively) at 1, 3 and 4 months after vaccination.

**Analytical approach:** We determined GMT and seroprotection and seroconversion rates. Chi-square or Fisher exact tests were used for categorical data; continuous values were analyzed using Wilcoxon rank-sum tests. Generalized estimating equation was used to determine association between GMT and age.

**Results:** 254 HD patients received HD-IIV3 (n=141), SD-IIV4 (n=36) or RIV4 (n=77) vaccine. A robust initial seroresponse to influenza A strains was observed after all 3 vaccines, with no difference in seroprotection rates at either the ≥1:40 or ≥1:160 titer at 1 and 2 months. Seroresponses to RIV4 and SD-IIV4 waned thereafter, such that by month 3 and 4, seroprotection by HD-IIV3 was significantly higher. Seroprotection rates were lower to the B strains across all three vaccines. Results trended similarly across patients aged below 65 years.

**Limitations:** Due to use of observational data, bias by unmeasured confounders may exist. Some of the subgroups by age were low in number.

**Conclusions:** Hemodialysis patients achieved high seroprotection rates after all 3 vaccines. The seroresponse waned more slowly with HD-IIV3 as compared to SD-IIV4 or RIV4 vaccine.

## Introduction

Influenza vaccination is the most effective method to prevent influenza infection and associated complications, including hospitalization and death. For various reasons, including impaired humoral and cell mediated immunity,^1,2^ close and frequent contact among patients in the outpatient dialysis clinic setting, and a high burden of comorbid illnesses, dialysis patients are susceptible to influenza infection and to severe morbidity, once infected. Beyond recommending vaccinating all dialysis patients and heeding caution with live vaccines, the Centers for Disease Control and Prevention (CDC) does not provide specific recommendations about vaccine type^3^. There are several vaccines available. These differ by 1) use of inactivated vs. recombinant virus; 2) the number of strains (3 in trivalent vs. 4 in quadrivalent); 3) the hemagglutinin dose per strain, (15 mcg in the standard dose inactivated vaccine vs. 45 mcg in high dose recombinant dose vs. 60 mcg in high dose inactivated); and 4) the use of adjuvants to augment the immune response.

Strong evidence supports use of high dose vaccine in the elderly general population. Serological studies show a nearly two fold increase in serotiters to the high dose (60 mcg per strain) as compared with standard dose (15 mcg /strain) inactivated vaccine^4^ and a landmark trial with over 30,000 subjects who were at least 65 years of age found a 24% reduction in laboratory confirmed influenza cases ^5^ and a 40% reduction in serious pneumonia.^6^ Greater efficacy of the high dose recombinant vaccine (45 mcg /strain) as compared with the standard dose (15 mcg /strain) inactivated vaccine has also been demonstrated. Specifically, the recombinant vaccine reduced influenza infections 30% as compared with the standard dose inactivated vaccine in a large randomized trial involving more than 9000 healthy subjects who were over 50 years old. A head-to-head comparison of the high dose recombinant and inactivated vaccine has not been done, for either seroresponse or clinical efficacy.

Moreover, none of these trials involved dialysis patients, who, in some observational studies, have been shown to generate a weakened and less durable seroresponse as compared with age-matched healthy controls. There have been two retrospective studies comparing clinical endpoints after receipt of the high and standard dose inactivated vaccine in dialysis populations, one based on an administrative database^7^ and the other, a national provider’s health information system^8^. These studies yielded conflicting results, and were also limited by selection bias (who received the high dose), and a lack of rigorously ascertained influenza-specific outcomes. To date, there has been no study comparing immunogenicity of the high dose vaccines with the standard dose vaccines in dialysis patients.

We leveraged a natural experiment to evaluate the seroresponse and its duration after vaccination with a high dose inactivated (HD-IIV3), a standard dose inactivated vaccine (SD-IIV4) and a high dose recombinant vaccine (RIV4) in a hemodialysis population. We were not able to evaluate seroresponse in patients administered influenza vaccine using adjuvants.

## Materials and Methods

### Study design

This is a prospective observational study that was conducted in the 2017-18 influenza season at four Dialysis Clinic Inc. hemodialysis clinics that employed different influenza vaccination strategies. Dialysis Clinic Inc. (DCI) is a national dialysis provider.

### Study Population

All patients at DCI clinics are offered an influenza vaccine annually. The medical director at each clinic decides which vaccines will be used at that site. The influenza vaccines that were on formulary within DCI in the 2017-2018 influenza season were: a high dose inactivated trivalent vaccine (HD-IIV3; Fluzone® High-Dose Influenza Vaccine, Sanofi Pasteur Inc.), a standard dose quadrivalent vaccine (SD-IIV4; Fluzone® Quadrivalent, Sanofi Pasteur Inc. and a high dose recombinant quadrivalent vaccine (RIV4; Flublok® Quadrivalent; Protein Sciences Inc.). The participating clinics and vaccines used at the clinic were as follows: DCI Boston: HD-IIV4 in all patients, DCI Walden Pond: SD-IIV3 in all patients, DCI Nashville: SD-IIV4 for patients less than 65 years of age and HD-IIV3 for patients 65 years old and over and DCI North Brunswick: RIV4 for all patients. The study population consisted of all hemodialysis patients who received an influenza vaccine at their dialysis clinic. Those who had received an influenza vaccine outside of their dialysis clinic were excluded. This quality improvement project was done to inform future vaccination policies at DCI, and the analytical files utilized de-identified data. DCI (staff unrelated to this project) kept a code to allow retrospective standard of care clinical data as described below to be linked to the hemagglutination inhibition (HI) titer data. A limited data set was created and a Data Use Agreement was executed between DCI and the researchers. The researchers did not have access to patient health information. The study was deemed exempt by the Western Institutional Review Board (File #1-1065223-1).

### Lab Samples

Pre-dialysis serum samples that were drawn monthly per standard of care were sent to the DCI Clinical Laboratory (Nashville, TN). The DCI Laboratory collected left over lab samples from these monthly routine blood draws. Excess serum (0.5 – 1 ml) were utilized to assess vaccination response prior to vaccination (baseline), 1 month, 3 month, and 4 months post vaccination. These samples were frozen at −80C and stored at the DCI Lab before being batch shipped at two intervals to the University of Wisconsin for HI titer assessment.

### Hemagglutination inhibition assay

Hemagglutination inhibition assays (HIA) for each virus strain were performed according to WHO standard microtiter methods.^9^ Laboratory staff performing the HIA were blinded to participant status. Briefly, the hemagglutinin antibody present in human sera was quantified based on the inhibition of virus-induced agglutination of guinea pig red blood cells (RBC).

Titrated influenza antigen was incubated with serially diluted sera for 30 min. Guinea pig RBCs were added and incubated for 60 minutes, and the dilution of serum that no longer inhibits hemagglutination was used as an index of antibody titer. Antibody concentrations below the lower limit of detection (< 1:10) were assigned a value of 1:1.

### Clinical Data

Information about influenza vaccination administered and the administration date in the current and prior seasons and demographic and clinical characteristics were extracted from DCI’s electronic health information system.

### Statistical Analyses

We first compared geometric mean HI antibody titers (GMT) for sera collected from all time points among HD-IIV3, SD-IIV4 and RIV4 groups. HI antibody titers at each time point were transformed to binary logarithms, and original values were divided by 4 (undetectable titer) to set the starting point of the log scale to zero prior to transformation. Average log_2_ titers at each of the 4 time points (baseline and approximately 1-month, 3-months, and 4-months post-vaccination) were calculated to obtain GMT over time.

The primary outcome measure was seroprotection defined by a HI titer ≥1:40.^10^ This concentration of antibody likely provides protection from infection at a rate of about 50% in immunocompetent individuals.^11,12^ Protection from infection improves with higher antibody concentrations, plateauing at approximately 1:160.^11–15^ Given the high risk of the dialysis population, and greater morbidity of the H1N1 and H3N2 strains,^16^ we evaluated seroprotection with an antibody HI titer ≥ 1:160 as a secondary outcome. This ≥1:160 threshold corresponds with protection of up to 95% patients.^11,13,15^

Additional secondary outcome measures included: (1) comparison of GMT of antibody and (2) seroconversion (defined as 4-fold or greater increase in baseline titer) rates. Categorical data were analyzed with an appropriate chi-square test or, when necessary, Fisher exact test; continuous values were analyzed using Wilcoxon rank-sum tests. Outcomes across vaccines were further compared by age subgroups (less than 65 years old vs. 65 years and over). Generalized estimating equation (GEE) was used to determine association between GMT and age. Statistical analyses were conducted using SAS 9.4.

## Results

Of 329 maintenance hemodialysis patients at the 4 participating clinics, 46 were excluded: 40 (12%) received vaccination elsewhere and 6 (2%) refused vaccination. Of the 283 vaccinated at participating clinics, 254 (90%) patients were present for all four follow-up months after vaccination. The vaccine administered was HD-IIV3 in 56% of patients, SD-IIV4 in 14% of patients, and RIV4 in 30% of patients. Mean age was 63 ± 14 years; 42% were female, median dialysis vintage was 46 ± 40 months, 40% were Black, and 35% had diabetes as cause of kidney failure. Baseline characteristics by vaccine type received are in Table 1. Patients receiving the SD-IIV4 were older than those receiving either the HD-IIV3 or RIV4-SD vaccine.

**Table 1.** Participant Characteristics at Baseline by vaccine type administered.

| **Characteristic** | **HD-IIV3**<br>**n=141** | **SD-IIV4**<br>**n=36** | **RIV4**<br>**n=77** | **Overall**<br>**n=254** |
| --- | --- | --- | --- | --- |
| **Age, years** | 62 (13) | 74 (125) | 62 (15) | 63 (14) |
| **% <65 years old** | 58% | 19% | 52% | 51% |
| **% Female** | 44% | 55% | 36% | 42% |
| **Race** |  |  |  |  |
| **White** | 14% | 91% | 33% | 31% |
| **Black** | 55% | 6% | 29% | 40% |
| **Others** | 20% | 3% | 16% | 16% |
| **Unknown** | 11% | 0% | 23 | 13% |
| **Ethnicity** |  |  |  |  |
| **Hispanic** | 4% | 3% | 4% | 4% |
| **Non-Hispanic** | 80% | 97% | 65% | 78% |
| **Unknown** | 16% | 0% | 31% | 18% |
| **ESKD Cause** |  |  |  |  |
| **Diabetes** | 34% | 17% | 46% | 35% |
| **Hypertension** | 28% | 17% | 20% | 24% |
| **Other/Unknown** | 38% | 67% | 35% | 41% |
| **Vintage, months** | 53 (46) | 29 (21) | 41 (34) | 46 (40) |
| **Albumin < 3.4 g/dL** | 5% | 3% | 3% | 4% |
| **Parathyroid hormone ≥ 600 pg/mL** | 30% | 16% | 13% | 23% |
| **Hemoglobin < 10 g/dL** | 18% | 6% | 10% | 14% |
| **Baseline Ferritin, ng/mL** | 1054 (437) | 1228 (452) | 1098 (411) | 1092 (435) |
| **KT/V <1.2** | 3% | 0% | 4% | 3% |
| **Urea Reduction Ratio %** | 74% (7%) | 75% (4%) | 74% (9%) | 74% (8%) |
| **Prior influenza vaccinations\*** | 7 (4) | 5 (2) | 7 (3) | 7 (4) |
| **Influenza vaccine received prior year** |  |  |  |  |
| **HD-IIV3** | 48% | 64% | 36% | 47% |
| **SD-IIV4** | 20% | 0% | 30% | 20% |
| **RIV4** | 0% | 0% | 0% | 0% |
| **Unknown type (e.g., received elsewhere)** | 24% | 36% | 33% | 28% |
| **Baseline use of Statin or Steroid** | 43% | 16% | 44% | 40% |
| **Hospitalization within 3 months PRIOR vaccination** | 23% | 19% | 10% | 19% |

Age, vintage, baseline ferritin, prior influenza vaccinations, and urea reduction ratio % are reported as mean (standard deviation). IIV3/IIV4 = egg-based trivalent/quadrivalent inactivated influenza vaccine; RIV4 = quadrivalent recombinant hemagglutinin influenza vaccine; HD =
High Dose trivalent inactivated influenza virus; SD-IIV4 = Standard Dose quadrivalent inactivated influenza virus; RIV4 = Recombinant influenza virus
\*number of prior influenza vaccinations recorded in dialysis facility electronic medical record.

### Serotiters

At 1 and 2 months post vaccination, GMT levels were higher against both H1N1 and H3N2 following vaccination with HD-IIV3 and RIV4 as compared with SD-IIV4 (**Table 2**). At 3 and 4 months, however, serotiters were significantly higher with HD-IIV3 as compared with both SD-IIV4 and RIV4. Serotiter values were lower across the board with all three vaccines for the B Strains (**Supplementary Table 1**) as compared with A strains. There was no difference in serotiters at any time point for the B/Victoria lineage by vaccine type. For the B/Yamagata lineage, which is not included in the HD-IIV3 but is in the other two vaccines, GMT levels were lower for the HD-IIV3 than the RIV4 at multiple time points. In the GEE model, all A-and B-viruses had lower GMT levels with advancing age however only GMT values against B-virus antigens were significant (mean difference −0.02 (95% CI: −0.03, −0.01; p< 0.001) B/Victoria and mean difference −0.01 (95% CI: −0.02, −0.00; p=0.01) B/Yamagata).

**Table 2.** Geometric mean titer (GMT) against influenza A virus (H1N1 and H3N2) by vaccine type and age group.

|  | GMT (95% Confidence Interval) |  |  | p-value |  |  |
| --- | --- | --- | --- | --- | --- | --- |
| **H1N1** | HD-IIV3 | SD-IIV4 | RIV4 | HD-IIV3<br>v. SD-IIV4 | HD-IIV3<br>v. RIV4 | RIV4 v.<br>SD-IIV4 |
| All Patients | 141 | 36 | 77 |  |  |  |
| Baseline | 147.2(128.4,168.6) | 111.0(87.2,141.2) | 151.6(128.5,178.8) | 0.06 | 0.79 | **0.03** |
| 1 mo | 324.8(278.8,378.3) | 163.1(116.8,227.8) | 369.6(288.1,474.2) | **0.0001** | 0.35 | **0.0002** |
| 2 mo | 284.4(250.1,323.4) | 217.7(167.9,282.3) | 287.2(221.1,373.1) | 0.06 | 0.94 | 0.13 |
| 3 mo | 290.0(251.0,335.1) | 145.3(108.5,194.6) | 134.8(89.6,203.0) | **<0.0001** | **0.0007** | 0.76 |
| 4 mo | 210.7(183.8,241.5) | 139.8(105.8,184.9) | 125.5(85.7,183.8) | **0.008** | **0.01** | 0.64 |
| <65 years old | N=82 | N=7 | N=40 |  |  |  |
| Baseline | 192.7(162.4,228.6) | 97.5(53.0,179.4) | 151.9(123.2,187.3) | **0.02** | 0.10 | 0.11 |
| 1 mo | 385.4(323.2,459.5) | 195.0(74.8,508.9) | 460.5(306.3,692.3) | **0.03** | 0.42 | 0.10 |
| 2 mo | 336.6(288.4,393.0) | 353.3(178.0,701.1) | 349.0(232.7,523.4) | 0.86 | 0.87 | 0.98 |
| 3 mo | 354.2(299.5,418.8) | 215.3(115.2,402.6) | 171.5(92.4,318.3) | 0.10 | **0.03** | 0.57 |
| 4 mo | 223.5(223.5,310.6) | 237.8(127.2,444.5) | 154.5(87.8,272.1) | 0.73 | 0.07 | 0.27 |
| ≥65 years old | N=59 | N=29 | N=37 |  |  |  |
| Baseline | 101.2(83.8,122.2) | 114.5(86.7,151.3) | 151.3(115.6,197.9) | 0.46 | **0.01** | 0.15 |
| 1 mo | 256.0(196.4,333.7) | 156.2(107.3,227.5) | 291.4(221.7,382.9) | **0.03** | 0.51 | **0.007** |
| 2 mo | 224.9(182.3,277.6) | 193.7(146.3,256.6) | 232.7(167.4,323.6) | 0.40 | 0.85 | 0.41 |
| 3 mo | 219.7(172.4,280.1) | 132.2(94.3,185.1) | 104.0(60.3,179.5) | **0.02** | **0.01** | 0.45 |
| 4 mo | 154.5(124.8,191.2) | 123.0(90.2,167.8) | 100.2(59.2,169.4) | 0.22 | 0.13 | 0.50 |
| **H3N2** |  |  |  |  |  |  |
| All Patients | 141 | 36 | 77 |  |  |  |
| Baseline | 134.0(114.2,157.4) | 77.0(56.6,104.6) | 141.1(112.6,176.6) | **0.002** | 0.71 | **0.002** |
| 1 mo | 274.8(230.6,327.3) | 127.0(83.5,193.2) | 350.1(268.8,456.1) | **0.0002** | 0.12 | **<0.0001** |
| 2 mo | 300.2(256.2,351.7) | 239.7(172.6,333.0) | 269.7(208.4,349.1) | 0.21 | 0.46 | 0.59 |
| 3 mo | 254.0(215.2,299.8) | 154.0(107.1,221.4) | 102.9(70.0,151.3) | **0.009** | **<0.0001** | 0.13 |
| 4 mo | 266.8(230.4,308.8) | 111.0(75.4,163.3) | 91.6(63.9,131.1) | **<0.0001** | **<0.0001** | 0.47 |
| <65 years old | N=82 | N=7 | N=40 |  |  |  |
| Baseline | 145.8(117.3,181.3) | 107.7(28.6,405.9) | 139.3(100.9,192.2) | 0.46 | 0.81 | 0.56 |
| 1 mo | 299.1(237.8,376.1) | 176.7(43.3,720.6) | 361.3(238.2,548.0) | 0.22 | 0.39 | 0.20 |
| 2 mo | 320.0(259.0,395.3) | 430.7(143.2,1295.8) | 269.1(182.5,396.7) | 0.44 | 0.39 | 0.35 |
| 3 mo | 270.2(216.6,337.2) | 237.8(70.2,805.0) | 102.0(59.6,174.5) | 0.75 | **0.001** | 0.21 |
| 4 mo | 294.1(242.3,356.8) | 176.7(41.3,755.9) | 90.3(55.2,147.7) | 0.43 | **<0.0001** | 0.30 |
| ≥65 years old | N=59 | N=29 | N=37 |  |  |  |
| Baseline | 119.3(93.8,151.6) | 71.0(53.5,94.2) | 143.0(102.9,198.7) | **0.01** | 0.36 | **0.002** |
| 1 mo | 244.2(185.2,322.1) | 117.3(74.9,183.7) | 338.5(241.5,474.4) | **0.004** | 0.14 | **0.0002** |
| 2 mo | 274.7(215.1,350.7) | 208.1(149.1,290.5) | 270.4(189.6,385.5) | 0.19 | 0.94 | 0.29 |
| 3 mo | 233.0(180.4,301.1) | 138.6(94.6,203.1) | 104.0(58.2,185.7) | **0.02** | **0.01** | 0.40 |
| 4 mo | 233.0(185.9,292.1) | 99.2(67.4,146.0) | 92.9(53.6,161.2) | **<0.0001** | **0.003** | 0.84 |

### Seroprotection

At one month post vaccination, seroprotection rates (≥1:40 titer) for the influenza A strains were high (greater than 90%) with all 3 vaccines (**Table 3, Figures 1A-B**). Significantly higher seroprotection rates were observed with HD-IIV3 for H1N1 and H3N2 as compared with RIV (E.g. 100 vs. 74% for both at month 4). Similarly, HD-IIV3 was associated with significantly higher seroprotection rates as compared with SD-IIV4 (100 vs. 56% for H1N1 and 100 vs. 85% for H3N2 at month 4). The same pattern of results is seen with seroprotection at the ≥1:160 titer level (**Table 3 and Figures 2A-B**), with HD-IIV3 achieving higher seroprotection rates at months 3 and 4 as compared with RIV4 and SD-IIV4. Similar results were observed in both age groups (less than 65 years old and 65 years and over) (**Supplementary Table 2, Supplementary Figure 1A-B and Supplementary Figure 2A-B.)**. For the B strains (**Table 3, Figures 1C-D**), no differences were seen in seroprotection rates by vaccine at any time point. This result also did not differ by age group (**Supplementary Table 2)**.

**Table 3.** Seroprotection rates (≥1:40 Influenza A and B virus and >1:160 A virus) against H1N1, H3N2, B/Victoria lineage and B/Yamagata lineage viruses by influenza vaccine type.

|  | % Seroprotected |  |  | p-value |  |  |
| --- | --- | --- | --- | --- | --- | --- |
|  | HD-IIV3<br>(n=141) | SD-IIV4<br>(n=36) | RIV4<br>(n=77) | HD-IIV3<br>v. SD-IIV4 | HD-IIV3 v.<br>RIV4 | RIV4 v. SD-IIV4 |
| **> 1:40** |  |  |  |  |  |  |
| **H1N1** |  |  |  |  |  |  |
| Baseline | 99.3 | 100.0 | 100.0 | 0.61 | 0.46 | 0.99 |
| 1 mo | 100.0 | 97.2 | 100.0 | **0.05** | 0.99 | 0.14 |
| 2 mo | 100.0 | 100.0 | 100.0 | 0.99 | 0.99 | 0.99 |
| 3 mo | 100.0 | 97.2 | 74.0 | **0.05** | **<.0001** | **0.003** |
| 4 mo | 100.0 | 55.6 | 74.0 | **0.05** | **<.0001** | **0.003** |
| **H3N2** |  |  |  |  |  |  |
| Baseline | 95.7 | 83.3 | 100.0 | **0.008** | 0.07 | **0.0002** |
| 1 mo | 99.3 | 91.7 | 100.0 | **0.006** | 0.46 | **0.01** |
| 2 mo | 100.0 | 100.0 | 100.0 | 0.99 | 0.99 | 0.99 |
| 3 mo | 99.3 | 91.7 | 72.7 | **0.006** | **<.0001** | **0.02** |
| 4 mo | 100.0 | 86.1 | 74.0 | **<.0001** | **<.0001** | 0.15 |
| **B/Victoria lineage** |  |  |  |  |  |  |
| Baseline | 38.3 | 38.9 | 32.5 | 0.95 | 0.39 | 0.50 |
| 1 mo | 67.4 | 58.3 | 64.9 | 0.31 | 0.72 | 0.50 |
| 2 mo | 68.1 | 63.9 | 61.0 | 0.63 | 0.30 | 0.77 |
| 3 mo | 60.3 | 63.9 | 57.1 | 0.69 | 0.65 | 0.50 |
| 4 mo | 49.3 | 52.8 | 56.0 | 0.71 | 0.35 | 0.75 |
| **B/Yamagata lineage** |  |  |  |  |  |  |
| Baseline | 30.5 | 25.0 | 33.8 | 0.52 | 0.62 | 0.35 |
| 1 mo | 45.4 | 38.9 | 58.4 | 0.48 | 0.07 | **0.05** |
| 2 mo | 42.6 | 33.3 | 49.4 | 0.32 | 0.33 | 0.11 |
| 3 mo | 29.8 | 36.1 | 36.4 | 0.46 | 0.32 | 0.98 |
| 4 mo | 30.0 | 30.6 | 36.0 | 0.95 | 0.37 | 0.57 |
| **> 1:160** |  |  |  |  |  |  |
| **H1N1** |  |  |  |  |  |  |
| Baseline | 61.0 | 50.0 | 63.6 | 0.23 | 0.70 | 0.17 |
| 1 mo | 90.1 | 72.2 | 92.2 | **0.005** | 0.60 | **0.005** |
| 2 mo | 88.7 | 80.6 | 79.2 | 0.20 | 0.06 | 0.87 |
| 3 mo | 87.9 | 63.9 | 61.0 | **0.001** | **<.0001** | 0.77 |
| 4 mo | 79.4 | 55.6 | 67.5 | **0.003** | **0.05** | 0.22 |
| **H3N2** |  |  |  |  |  |  |
| Baseline | 56.7 | 30.6 | 51.9 | **0.005** | 0.50 | **0.03** |
| 1 mo | 77.3 | 52.8 | 83.1 | **0.003** | 0.31 | **0.001** |
| 2 mo | 85.8 | 80.6 | 76.6 | 0.43 | 0.09 | 0.64 |
| 3 mo | 76.6 | 63.9 | 54.5 | 0.12 | **0.0008** | 0.35 |
| 4 mo | 83.0 | 50.0 | 50.6 | **<.0001** | **<.0001** | 0.95 |

For comparisons that were identical, p-value assigned was 0.99.

**Figure 1.**
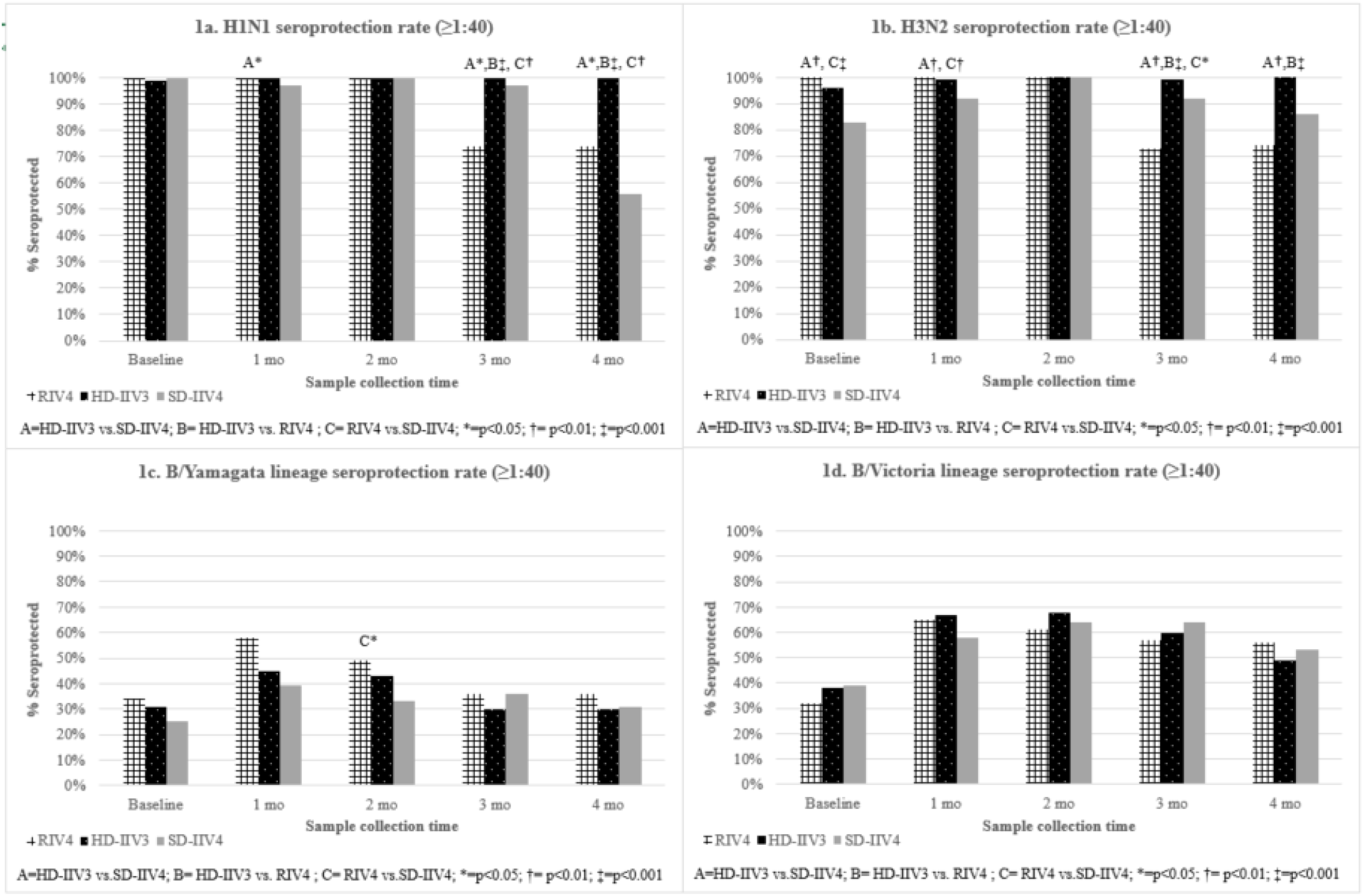
**Seroprotection frequency, defined by hemagglutination inhibition (HI) titer >1:40 for influenza A and B virus after immunization with the RIV4-SD, HD-IIV3 or SD-IIV4-SD vaccine in hemodialysis patients**.

**Figure 2.**
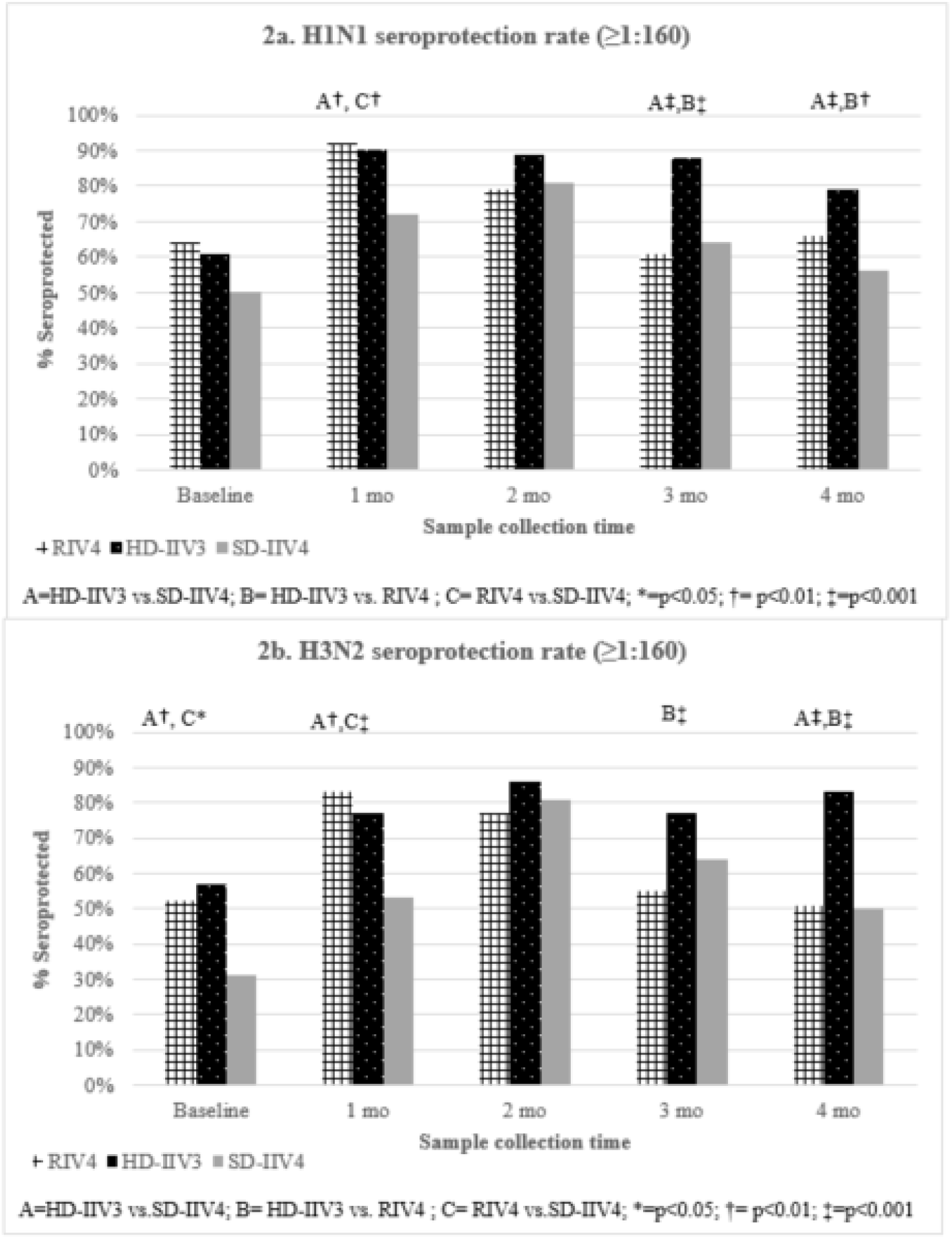
**Seroprotection frequency, defined by hemagglutination inhibition (HI) titer ≥1:160 for influenza A (H1N1 and H3N2) after immunization with RIV4-SD, HD-IIV3 or SD-IIV4-SD vaccine in hemodialysis patients**.

Comparisons of seroprotection rates after 4 months follow-up between age groups and vaccine type are shown in **Supplementary Figures 1 and 2**. At the ≥ 1:40 threshold, seroprotection rates were generally higher in younger patients compared to older patients, but the difference was statistically significant in only seroprotection against the B/Victoria strain among HD-IIV3 recipients. At the ≥1:160 threshold, the difference by age group was statistically significant against H1N1 among HD-IIV3 and RIV vaccine recipients.

### Seroconversion rates

Seroconversion occurred more frequently among patients receiving the HD-IIV3 or RIV4 vaccines against the influenza A strains (**Supplementary Table 3**), although there was considerable month-to-month variability. Seroconversion versus influenza B strains were uniformly low. Seroconversion rates observed in patients less than 65 and those 65 years of age and older against influenza A and B viruses are provided in **Supplement Table 4**.

## Discussion

Hemodialysis patients in both age groups generated a robust initial seroresponse to each of the three seasonal influenza vaccines tested, with achievement of serotiters that are well within the range of those reported in the age-matched general population.^4,17,18^ This study, however, identifies an important difference between the vaccines in the durability of the response elicited. Over a 4 month period, we find slower waning of the seroresponse with HD-IIV3 as compared with RIV4 and SD-IIV4. At the end of follow up, serotiters for influenza A strains were more than twice as high after HD-IIV3 than after RIV4 and SD-IIV4 and seroprotection (at both ≥1:40 and at ≥1:160 titers) was significantly more likely. This result was consistent in both age groups.

Of note, there were no differences among vaccines at any time point in seroresponse to B strains, including for the B/Yamagata strain which is the 4^th^ virus included in the quadrivalent but not the trivalent vaccine. This is consistent with previous findings of cross-reactivity against influenza B virus.^19^ In a paired pre- and post-vaccination study serum samples found antibodies to HA from both B/Victoria- or B/Yamagata-lineage, even in subjects who received IIV3 vaccine (which did not contain HA from the B/Victoria lineage). Influenza B virus cross-reactive memory B cells are common in humans and express both neutralizing and non-neutralizing immunoglobulins.^20^

There are several studies showing the greater efficacy and immunogenicity in flu vaccines that contain a greater quantity of HA antigen, that is, RIV4 and HD-IIV3. Therefore the greater durability of response in the HD-IIV3 group in our study is consistent with the greater efficacy and immunogenicity found in other studies, which have not reported titers more than 1 month post-vaccination.^5,6,21^

In a randomized, double-blind, multicenter trial comparing RIV4 and SD-IIV4 in adults 50 years of age or older, RIV4 was shown to be superior to SD-IIV4 against confirmed influenza like illness.^21^ In our study, patients 65 years or older vaccinated with RIV4 influenza vaccine had higher GMT values and seroprotection rates at ≥1:40 and ≥1:160 levels against H1N1 and H3N2 post vaccination when compared to patients vaccinated with SD-IIV4 influenza vaccine. These differences were not observed in patients less than 65 years old. Our findings suggest that RIV4 may be superior to SD-IIV4 in hemodialysis patients and warrants further investigation.

There are no randomized clinical trials comparing vaccine efficacy among HD-IIV3, SD-IIV4 or RIV4 in dialysis patients. Among patients 65 years of age and older, DiazGranados et al report GMT values against H1N1 with HD-IIV3 were 1.8 times higher than that observed with SD-IIV3 when measured 1 month post vaccination.^6^ In our study, hemodialysis patients GMT values against H1N1 with HD-IIV3 were 1.6 times higher in patients 65 years of age and older and two times higher in patients less than 65 years old than that observed with SD-IIV3 when measured 1 month post vaccination. Unfortunately, DiazGranados et al do not report serotiters after 1 month. Our study is first to report GMT values over 4 consecutive months in patients requiring hemodialysis, illustrating slower waning of the high dose vaccine. Additionally, our seroresponse data predicts greater clinical efficacy with high dose vaccine and likely a reason for greater clinical efficacy in observational trial.^8^ However, further investigation is warranted.

There have been no prior comparisons of the RIV4 with HD-IIV3 for seroresponse or clinical outcomes in dialysis or, for that matter, the general population. While the RIV4 can be produced in large quantities more readily than the HD-IIV3, our seroresponse data suggests quicker waning and significantly lower seroprotection at 3-4 months. This may be particularly important in a population with high risk for influenza related morbidity, as is the case for a dialysis population. Clinical trials comparing influenza vaccines with or without neuraminidase component are needed in the dialysis population.

The excellent seroresponse seen in this paper, even with the SD-IIV4, contrasts with some (but not all) prior studies that have reported poor seroresponse to influenza vaccine in dialysis patients. It is expected that seroprotection rates would decrease over time after vaccination, as seen in Table 3. Our results are similar to a cohort study of 42 Thai patients utilizing hemodialysis that received a single standard dose of trivalent influenza vaccine containing 15mcg of H1N1, H3N2 and B-Victoria strains during the 2016-2017 influenza season.^22^ Nongnuch et al reported seroprotection (≥1:40) to combination of each strain as 35.7%, 71.4%, 61.9% and 47.6% at baseline, 1 month, 6 months and 12 months, respectively. Applying the same methodology to our cohort, the percent of patients exhibiting seroprotection to all three H1N1, H3N2, and B-Victoria strains would be 32.5% and 58.3% at baseline and 1 month post vaccination, respectively. Additionally a Taiwanese study found seroprotection (≥1:40) rates for H1N1 at 1 month after receipt of a standard dose inactivated vaccine by age group (less than 65 and 65 years of age or older) of 43% and 38% respectively.^23^ Seroprotection rates in hemodialysis patients after receipt of the 2009 pandemic influenza vaccine were 57% in one study^24^ and 33%^25^ in another. In contrast, another study of the same vaccine found seroprotection rates of 50% at 4 weeks and 39% at 24 weeks.^26^

Uremia and anemia have been shown to be associated with variation in the immune response to influenza vaccine ^27^, but would not explain results here given no difference in these parameters at baseline (Table 1). The more likely explanation for the very robust seroresponse in the present study is the high rate of prior vaccination and that the circulating H1N1 strain has been relatively constant since 2010.^28,29^ Previous experience with influenza vaccine affects the immune response to the current vaccine.^30^ The study cohort has an extensive influenza vaccine history. The vaccine composition was very similar to the 2016-17 vaccine. Also, the pandemic H1N1 virus has been included since 2010. Although the H1N1 virus did change in 2017, the two vaccine viruses, A/California and A/Michigan, are antigenically similar with the primary differences in the neuraminidase component which is not measured by hemagglutination inhibition.^28^ Many studies show relatively high baseline antibody concentrations and seroprotection with the 2017-18 influenza vaccine.^31–34^ No study that we found has a study population with an immunization history that matches ours.

The immunologic memory (both humoral and cell-mediated) induced by prior vaccination enhances the seroresponse, thereby reducing influenza cases and morbidity. This is likely true for previously seen epitopes, however, there is a body of literature that suggests that, via epitope masking, the cross-reactive antibodies may inhibit the response to novel epitopes.^35^ Accordingly it is possible that a greater and more durable seroresponse may be a disadvantage in the setting of a major antigen drift. The role of pre-existing immunity, of harm or benefit in the setting of major antigenic drifts, remains controversial and is the subject of ongoing research.^36^

There are some limitations to our study. First, our results were based upon data obtained from routine clinical practice and thus, not based upon a prospective randomized controlled trial. There may be unmeasured confounders biasing our results such as the important differences in baseline characteristics (e.g., age, race) seen among groups. Selection bias by vaccine type may have played a role, though we believe that the potential for selection bias is minimized given that the vaccine type administered was a decision made at the clinic as opposed to at the patient level. Furthermore, we do not know if our patients experienced influenza during our study period, and whether this, (if not equally distributed across vaccine types) rather than the vaccination, affected serotiters. Additionally, the mean age in SD-IIV4 group was significantly higher than the other two groups, and older age may blunt patient seroresponse. When patients were divided into age groups, some of the comparison groups were quite small, reducing precision of estimates within each vaccine type. Nonetheless, serotiters were consistently higher in patients less than 65 years old compared to those 65 years of age and older at each time period (Table 2 and Figures 1–2).

In summary, our study finds an excellent seroresponse to all influenza vaccines used in this population, higher than previously reported, which may be a result of high rates of prior year vaccination and minimal antigenic drift in influenza strains in recent years. The key difference among vaccines is a slower waning of HD-IIV3 over time such that by 3 and 4 months, seroprotection rates were significantly greater as compared with RIV4 and SD-IIV4. In our limited sample, the effect pattern was similar by age. These results are consistent with a large randomized controlled trial showing a reduction in influenza cases with HD-IIV3 over SD-IIV3 in the elderly. Our results could be used to support the prudent option to recommend HD-IIV3 over SD-IIV4 in dialysis patients when the choice is available, with supplies prioritized for elderly patients when needed, given their lower seroresponse and higher risk for morbidity.

## Data Availability

All data in the present study are available upon reasonable request to the authors

## Author contributions

Research idea and study design: HJM, EKL, GA, DEW, DCM, TK, KBM, DSJ; data acquisition: HJM, GA, MSH; data analysis/interpretation: HJM, GA, MSH, DEW, DCM, CMH, EKL. Each author contributed important intellectual content during manuscript drafting or revision, accepts personal accountability for the author’s own contributions, and agrees to ensure that questions pertaining to the accuracy or integrity of any portion of the work are appropriately investigated and resolved.

## Supplemental Material Table of Contents

Supplement Table 1. Geometric mean titer (GMT) results against influenza B virus (B/Victoria lineage and B/Yamagata lineage) by vaccine type and age group.

Supplement Table 2. Seroprotection rates (≥1:40 Influenza A and B virus and >1:160 A virus) against H1N1, H3N2, B/Victoria lineage and B/Yamagata lineage viruses by influenza vaccine type and age group.

Supplement Table 3. Seroconversion rates (≥ 4x baseline titer) against H1N1, H3N2, B/Victoria lineage and B/Yamagata lineage viruses by influenza vaccine type.

Supplement Table 4. Seroconversion rates (≥ 4x baseline titer) against H1N1, H3N2, B/Victoria lineage and B/Yamagata lineage viruses by influenza vaccine type and age group.

Supplement Figure 1. Seroprotection rates (hemagglutination inhibition titer ≥1:40) between age groups for each influenza virus and influenza vaccine type.

Supplement Figure 2. Seroprotection rates (hemagglutination inhibition titer ≥1:160) between age groups for each influenza virus and influenza vaccine type.

