## supplemental tables for "Seroresponse to Inactivated and Recombinant Influenza Vaccines among Maintenance Hemodialysis Patients"

**Supplement**

**Supplement Table 1. Geometric mean titer (GMT) results against influenza B virus (B/Victoria lineage and B/Yamagata lineage) by vaccine type and age group.**

| | GMT (95% Confidence Interval) | | | p-value | | |
|---|---|---|---|---|---|---|
| B/Victoria lineage | HD-IIV3 | SD-IIV4 | RIV4 | HD-IIV3 v. SD-IIV4 | HD-IIV3 v. RIV4 | RIV4 v. SD-IIV4 |
| All Patients | N=141 | N=36 | N=77 | | | |
| Baseline | 26.6(23.5,30.1) | 25.7(19.7,33.4) | 23.5(19.6,28.2) | 0.80 | 0.26 | 0.58 |
| 1 mo | 47.0(40.4,54.8) | 40.8(29.6,56.1) | 46.6(37.3,58.3) | 0.41 | 0.95 | 0.50 |
| 2 mo | 46.8(40.7,53.9) | 44.9(32.9,61.2) | 43.4(34.2,55.0) | 0.79 | 0.58 | 0.87 |
| 3 mo | 38.5(32.7,45.2) | 44.0(31.9,60.8) | 40.4(31.8,51.2) | 0.45 | 0.73 | 0.67 |
| 4 mo | 33.0(28.4,38.3) | 33.0(24.2,45.0) | 37.1(29.7,46.4) | 1.0 | 0.37 | 0.54 |
| <65 years old | N=82 | N=7 | N=40 | | | |
| Baseline | 34.1(28.7,40.4) | 32.8(16.1,67.0) | 24.2(18.7,32.3) | 0.90 | **0.03** | 0.36 |
| 1 mo | 65.3(53.7,79.5) | 44.2(20.3,96.2) | 60.6(44.0,83.5) | 0.27 | 0.68 | 0.44 |
| 2 mo | 61.0(51.1,73.0) | 59.4(28.7,123.0) | 53.7(37.5,76.8) | 0.93 | 0.52 | 0.82 |
| 3 mo | 49.4(40.2,60.7) | 59.4(22.6,156.7) | 48.4(34.1,60.8) | 0.62 | 0.91 | 0.65 |
| 4 mo | 43.9(36.1,53.5) | 36.2(13.2,99.3) | 42.9(30.4,60.6) | 0.59 | 0.90 | 0.70 |
| ≥65 years old | N=59 | N=29 | N=37 | | | |
| Baseline | 18.9(16.3,21.8) | 24.2(18.0,32.6) | 22.8(17.5,29.7) | 0.13 | 0.21 | 0.76 |
| 1 mo | 29.8(24.5,36.3) | 40.0(27.6,58.1) | 35.1(26.1,47.2) | 0.12 | 0.34 | 0.57 |
| 2 mo | 32.4(26.7,39.3) | 42.0(29.3,60.1) | 34.4(25.4,46.7) | 0.16 | 0.72 | 0.39 |
| 3 mo | 27.1(21.4,34.4) | 41.0(28.7,58.5) | 33.2(24.1,45.7) | **0.05** | 0.31 | 0.37 |
| 4 mo | 22.2(18.2,27.1) | 32.3(23.0,45.3) | 31.7(23.9,42.3) | **0.04** | **0.04** | 0.94 |
| B/Yamagata lineage | HD-IIV3 | SD-IIV4 | RIV4 | HD-IIV3 v. SD-IIV4 | HD-IIV3 v. RIV4 | RIV4 v. SD-IIV4 |
| All Patients | N=141 | N=36 | N=77 | | | |
| Baseline | 21.4(19.1,24.0) | 18.9(14.2,25.0) | 25.7(20.9,31.7) | 0.35 | 0.13 | 0.09 |
| 1 mo | 27.9(24.3,32.0) | 25.7(18.9,34.9) | 40.0(32.6,49.1) | 0.59 | **0.003** | **0.02** |
| 2 mo | 27.4(24.2,31.0) | 24.2(18.4,32.0) | 36.6(29.3,45.6) | 0.39 | **0.03** | **0.03** |
| 3 mo | 21.1(18.4,24.3) | 22.4(16.2,31.2) | 26.2(21.2,32.4) | 0.70 | 0.08 | 0.42 |
| 4 mo | 20.6(18.0,23.5) | 20.0(14.9,26.9) | 27.6(22.3,34.3) | 0.85 | **0.02** | 0.09 |
| <65 years old | N=82 | N=7 | N=40 | | | |
| Baseline | 24.9(21.4,29.0) | 26.9(11.9,60.9) | 29.3(22.4,38.4) | 0.78 | 0.26 | 0.81 |
| 1 mo | 33.5(28.1,39.9) | 36.2(16.6,78.9) | 47.6(36.5,62.0) | 0.80 | **0.03** | 0.43 |
| 2 mo | 30.8(26.2,36.1) | 36.2(16.6,78.9) | 45.2(33.6,60.7) | 0.58 | **0.01** | 0.56 |
| 3 mo | 23.7(19.8,28.3) | 40.0(13.2,121.4) | 31.4(23.6,41.8) | 0.12 | 0.08 | 0.53 |
| 4 mo | 23.3(19.6,27.7) | 29.7(11.3,78.3) | 32.3(24.1,43.3) | 0.45 | **0.04** | 0.83 |
| ≥65 years old | N=59 | N=29 | N=37 | | | |
| Baseline | 17.4(14.7,20.5) | 17.3(12.7,23.6) | 22.4(16.2,31.0) | 0.99 | 0.17 | 0.26 |
| 1 mo | 21.7(17.8,26.6) | 23.6(16.7,33.5) | 33.2(24.1,45.7) | 0.65 | **0.02** | 0.15 |
| 2 mo | 23.3(19.2,28.3) | 22.0(16.2,29.9) | 29.1(20.9,40.4) | 0.74 | 0.22 | 0.22 |
| 3 mo | 18.0(14.4,22.5) | 19.5(14.0,27.3) | 21.6(15.7,29.6) | 0.68 | 0.34 | 0.67 |
| 4 mo | 17.4(14.1,21.3) | 18.2(13.3,24.9) | 23.3(16.8,32.4) | 0.80 | 0.10 | 0.27 |

**Supplement Table 2. Seroprotection rates (≥1:40 Influenza A and B virus and >1:160 A virus) against H1N1, H3N2, B/Victoria lineage and B/Yamagata lineage viruses by influenza vaccine type and age group.**

| | < 65 years old | | | | | | ≥65 years old | | | | | |
|---|---|---|---|---|---|---|---|---|---|---|---|---|
| | % Seroprotected | | | p-value | | | % Seroprotected | | | p-value | | |
| | HD-IIV3 (n=82) | SD-IIV4 (n=7) | RIV4 (n=40) | HD-IIV3 v. SD-IIV4 | HD-IIV3 v. RIV4 | RIV4 v. SD-IIV4 | HD-IIV3 (n=59) | SD-IIV4 (n=29) | RIV4 (n=37) | HD-IIV3 v. SD-IIV4 | HD-IIV3 v. RIV4 | RIV4 v. SD-IIV4 |
| **> 1:40** | | | | | | | | | | | | |
| **H1N1** | | | | | | | | | | | | |
| Baseline | 100 | 100 | 100 | 0.99 | 0.99 | 0.99 | 98 | 100 | 100 | 0.48 | 0.99 | 0.99 |
| 1 mo | 100 | 100 | 100 | 0.99 | 0.99 | 0.99 | 100 | 97 | 100 | 0.15 | 0.43 | 0.26 |
| 2 mo | 100 | 100 | 100 | 0.99 | 0.99 | 0.99 | 100 | 100 | 100 | 0.99 | 0.99 | 0.99 |
| 3 mo | 100 | 100 | 75 | 0.99 | **<0.001** | 0.14 | 100 | 97 | 73 | 0.15 | **<0.001** | **0.01** |
| 4 mo | 100 | 100 | 75 | 0.99 | **<0.001** | 0.14 | 100 | 97 | 73 | 0.15 | **<0.001** | **0.01** |
| **H3N2** | | | | | | | | | | | | |
| Baseline | 96 | 71 | 100 | **0.006** | 0.22 | **<0.001** | 95 | 86 | 100 | 0.16 | 0.16 | **0.02** |
| 1 mo | 100 | 100 | 100 | 0.99 | 0.99 | 0.99 | 98 | 90 | 100 | 0.07 | 0.43 | **0.05** |
| 2 mo | 100 | 100 | 100 | 0.99 | 0.99 | 0.99 | 100 | 100 | 100 | 0.99 | 0.99 | 0.99 |
| 3 mo | 99 | 100 | 75 | 0.77 | **<0.001** | 0.14 | 100 | 90 | 70 | **0.01** | **<0.001** | 0.06 |
| 4 mo | 100 | 86 | 75 | **<0.001** | **<0.001** | 0.54 | 100 | 86 | 73 | **0.004** | **<0.001** | 0.19 |
| **B/Victoria lineage** | | | | | | | | | | | | |
| Baseline | 54 | 57 | 33 | 0.86 | 0.03 | 0.21 | 17 | 35 | 32 | 0.07 | 0.08 | 0.86 |
| 1 mo | 82 | 57 | 70 | 0.12 | 0.14 | 0.50 | 48 | 59 | 60 | 0.32 | 0.25 | 0.95 |
| 2 mo | 83 | 86 | 70 | 0.85 | 0.10 | 0.39 | 49 | 59 | 51 | 0.32 | 0.71 | 0.56 |
| 3 mo | 70 | 71 | 63 | 0.91 | 0.44 | 0.65 | 49 | 62 | 51 | 0.20 | 0.71 | 0.38 |
| 4 mo | 63 | 57 | 62 | 0.76 | 0.88 | 0.82 | 31 | 52 | 50 | **0.05** | 0.06 | 0.89 |
| **B/Yamagata lineage** | | | | | | | | | | | | |
| Baseline | 39 | 43 | 40 | 0.84 | 0.92 | 0.89 | 19 | 21 | 27 | 0.81 | 0.33 | 0.55 |
| 1 mo | 57 | 57 | 68 | 0.99 | 0.28 | 0.59 | 29 | 35 | 49 | 0.59 | 0.05 | 0.25 |
| 2 mo | 51 | 57 | 63 | 0.76 | 0.24 | 0.79 | 31 | 28 | 35 | 0.78 | 0.64 | 0.51 |
| 3 mo | 37 | 57 | 45 | 0.28 | 0.37 | 0.55 | 20 | 31 | 27 | 0.27 | 0.45 | 0.72 |
| 4 mo | 36 | 57 | 44 | 0.26 | 0.41 | 0.51 | 22 | 24 | 28 | 0.82 | 0.53 | 0.74 |
| **> 1:160** | | | | | | | | | | | | |
| **H1N1** | | | | | | | | | | | | |

| | | | | | | | | | | | | |
|---|---|---|---|---|---|---|---|---|---|---|---|---|
| Baseline | 74 | 29 | 65 | 0.01 | 0.28 | 0.07 | 42 | 55 | 62 | 0.26 | 0.06 | 0.57 |
| 1 mo | 96 | 86 | 94 | 0.19 | 0.36 | 0.55 | 81 | 69 | 92 | 0.19 | 0.15 | **0.02** |
| 2 mo | 94 | 100 | 80 | 0.50 | 0.02 | 0.19 | 81 | 76 | 78 | 0.55 | 0.72 | 0.81 |
| 3 mo | 96 | 86 | 68 | 0.19 | **<0.001** | 0.33 | 76 | 59 | 54 | 0.09 | **0.02** | 0.71 |
| 4 mo | 89 | 86 | 75 | 0.79 | **0.04** | 0.54 | 66 | 48 | 60 | 0.11 | 0.51 | 0.37 |
| **H3N2** | | | | | | | | | | | | |
| Baseline | 57 | 43 | 48 | 0.46 | 0.31 | 0.82 | 56 | 28 | 57 | **0.01** | 0.94 | **0.02** |
| 1 mo | 79 | 57 | 78 | 0.18 | 0.82 | 0.25 | 75 | 52 | 89 | **0.03** | 0.08 | **0.001** |
| 2 mo | 83 | 86 | 75 | 0.85 | 0.30 | 0.54 | 90 | 79 | 78 | 0.18 | 0.12 | 0.93 |
| 3 mo | 77 | 57 | 53 | 0.25 | **0.006** | 0.82 | 76 | 66 | 57 | 0.29 | **0.04** | 0.47 |
| 4 mo | 84 | 57 | 53 | 0.07 | **<0.001** | 0.82 | 81 | 48 | 49 | **0.001** | **0.001** | 0.98 |

For comparisons that were identical, p-value assigned was 0.99.

**Supplement Table 3. Seroconversion rates (≥ 4x baseline titer) against H1N1, H3N2, B/Victoria lineage and B/Yamagata lineage viruses by influenza vaccine type.**

| | % Seroconverted | | | p-value | | |
|---|---|---|---|---|---|---|
| | HD-IIV3 (n=141) | SD-IIV4 (n=36) | RIV4 (n=77) | HD-IIV3 v. SD-IIV4 | HD-IIV3 v. RIV4 | RIV4 v. SD-IIV4 |
| **H1N1** | | | | | | |
| 1 mo | 34.0 | 13.9 | 35.1 | **0.02** | 0.88 | **0.02** |
| 2 mo | 24.8 | 33.3 | 28.6 | 0.30 | 0.55 | 0.61 |
| 3 mo | 29.8 | 8.3 | 24.7 | **0.008** | 0.42 | 0.04 |
| 4 mo | 14.9 | 11.1 | 18.2 | 0.56 | 0.53 | 0.34 |
| **H3N2** | | | | | | |
| 1 mo | 31.9 | 16.2 | 42.9 | 0.07 | 0.11 | **0.006** |
| 2 mo | 34.0 | 51.4 | 31.2 | 0.08 | 0.67 | **0.03** |
| 3 mo | 23.4 | 24.3 | 16.9 | 0.84 | 0.26 | 0.37 |
| 4 mo | 29.8 | 13.5 | 13.0 | **0.05** | **0.005** | 0.99 |
| **B/Victoria** | | | | | | |
| 1 mo | 19.9 | 13.9 | 26.0 | 0.41 | 0.30 | 0.15 |
| 2 mo | 16.3 | 11.1 | 19.5 | 0.44 | 0.56 | 0.27 |
| 3 mo | 13.5 | 13.9 | 19.5 | 0.95 | 0.24 | 0.47 |
| 4 mo | 7.1 | 5.6 | 14.7 | 0.74 | 0.08 | 0.16 |
| **B/Yamagata** | | | | | | |
| 1 mo | 6.4 | 5.6 | 18.2 | 0.85 | **0.006** | 0.07 |
| 2 mo | 3.5 | 2.8 | 14.3 | 0.82 | **0.003** | 0.06 |
| 3 mo | 2.8 | 5.6 | 3.9 | 0.42 | 0.67 | 0.69 |
| 4 mo | 2.1 | 0 | 6.7 | 0.38 | 0.09 | 0.11 |

**Supplement Table 4. Seroconversion rates (≥ 4x baseline titer) against H1N1, H3N2, B/Victoria lineage and B/Yamagata lineage viruses by influenza vaccine type and age group.**

| | < 65 years old | | | | | | ≥65 years old | | | | | |
|---|---|---|---|---|---|---|---|---|---|---|---|---|
| | % Seroconverted | | | p-value | | | % Seroconverted | | | p-value | | |
| | HD-IIV3 (n=82) | SD-IIV4 (n=7) | RIV4 (n=40) | HD-IIV3 v. SD-IIV4 | HD-IIV3 v. RIV4 | RIV4 v. SD-IIV4 | HD-IIV3 (n=59) | SD-IIV4 (n=29) | RIV4 (n=37) | HD-IIV3 v. SD-IIV4 | HD-IIV3 v. RIV4 | RIV4 v. SD-IIV4 |
| **H1N1** | | | | | | | | | | | | |
| 1 mo | 32 | 29 | 50 | 0.86 | **0.05** | 0.29 | 37 | 10 | 19 | **0.008** | 0.06 | 0.33 |
| 2 mo | 22 | 86 | 43 | **0.0003** | **0.02** | **0.03** | 29 | 21 | 14 | 0.41 | 0.08 | 0.44 |
| 3 mo | 27 | 14 | 35 | 0.47 | 0.35 | 0.28 | 34 | 7 | 14 | **0.006** | **0.03** | 0.39 |
| 4 mo | 9 | 29 | 28 | 0.09 | **0.006** | 0.95 | 24 | 7 | 8. | **0.05** | **0.05** | 0.85 |
| **H3N2** | | | | | | | | | | | | |
| 1 mo | 32 | 14 | 48 | 0.46 | 0.09 | 0.10 | 32 | 17.2 | 38 | 0.14 | 0.5716 | 0.07 |
| 2 mo | 32 | 86 | 35 | 0.34 | 0.72 | **0.01** | 37 | 41.4 | 27 | 0.71 | 0.30 | 0.22 |
| 3 mo | 23 | 43 | 15 | **0.004** | 0.29 | 0.08 | 24 | 20.7 | 19 | 0.75 | 0.58 | 0.86 |
| 4 mo | 32 | 29 | 13 | 0.25 | **0.02** | 0.27 | 27 | 10.3 | 14 | 0.07 | 0.11 | 0.70 |
| **B/Victoria** | | | | | | | | | | | | |
| 1 mo | 22 | 14 | 38 | 0.63 | 0.07 | 0.23 | 17 | 14 | 1 | 0.71 | 0.65 | 0.97 |
| 2 mo | 15 | 14 | 28 | 0.98 | 0.09 | 0.46 | 19 | 11 | 11 | 0.32 | 0.30 | 0.95 |
| 3 mo | 11 | 14 | 28 | 0.79 | **0.02** | 0.46 | 17 | 11 | 11 | 0.70 | 0.41 | 0.71 |
| 4 mo | 6 | 14 | 21 | 0.41 | **0.02** | 0.70 | 9 | 8 | 8 | 0.38 | 0.98 | 0.42 |
| **B/Yamagata** | | | | | | | | | | | | |
| 1 mo | 5 | 0 | 25 | 0.55 | **0.001** | 0.14 | 9 | 7 | 11 | 0.80 | 0.70 | 0.58 |
| 2 mo | 1 | 0 | 23 | 0.77 | **<0.0001** | 0.16 | 7 | 3 | 5 | 0.53 | 0.79 | 0.71 |
| 3 mo | 0 | 14 | 8 | **0.0006** | **0.01** | 0.55 | 7 | 3 | 0 | 0.53 | 0.11 | 0.26 |
| 4 mo | 0 | 0 | 10 | 0.99 | **0.003** | 0.38 | 5 | 0 | 3 | 0.22 | 0.59 | 0.37 |
